# Understanding technology-related prescribing errors for system optimisation: the Technology-Related Error Mechanism (TREM) classification

**DOI:** 10.1101/2024.09.02.24312874

**Authors:** Magdalena Z. Raban, Alison Merchant, Erin Fitzpatrick, Melissa T. Baysari, Ling Li, Peter J. Gates, Johanna I. Westbrook

**Author notes:** Corresponding author: Dr Magda Raban, E, Level 6, 75 Talavera Rd Macquarie Park NSW 2109. MZR and AM are joint first authors on this manuscript.

## Abstract

**Objectives:** Technology-related prescribing errors curtail the positive impacts of computerised provider order entry (CPOE) on medication safety. Understanding how technology-related errors occur can inform CPOE optimisation. Previously, we developed a classification of the underlying mechanisms of technology-related errors using prescribing error data from two adult hospitals. Our objective was to update the classification using paediatric prescribing error data, and to assess the reliability with which reviewers could independently apply the classification.

**Materials and Methods:** Using data on 1696 prescribing errors identified by chart review in 2016 and 2017 at a tertiary paediatric hospital, we identified errors that were technology-related. These errors were investigated to classify their underlying mechanisms using our previously developed classification, and new categories were added based on the data. A two-step process was used to identify and classify technology-related errors involving review of the error in the CPOE and simulating the error in the CPOE testing environment.

**Results:** The Technology-Related Error Mechanism (TREM) classification comprises seven categories and 19 subcategories. The seven categories are: 1) errors due to incorrect system configuration or system malfunction, 2) prescribing on the wrong patient record, 3) selection errors, 4) construction errors, 5) editing errors, 6) errors that occur when using workflows that differ from a paper-based system 7) contributing factor: use of hybrid systems.

**Conclusion:** Technology-related errors remain a critical issue for CPOE. The updated TREM classification provides a systematic means of assessing and monitoring technology-related errors to inform and prioritise system improvements, and has now been updated for the paediatric setting.

**What is already known on this topic:**

- Technology-related errors occur frequently in computerised provider order entry (CPOE).
- Technology-related errors can be addressed by CPOE modifications; however, an understanding of how the errors occurred is required.

**What this study adds:**

- This study presents a method for classifying how technology-related errors occur, the Technology-Related Error Mechanism (TREM) classification.

*How this study might affect research, practice or policy:*

- Using the TREM classification can aid system managers in identifying areas for CPOE optimisation to deliver improved patient safety outcomes.

## INTRODUCTION

Computerised provider order entry (CPOE) is a computer-based system for placing orders (e.g. medications, pathology tests, imaging, blood products) used in hospitals and now most commonly integrated into an electronic medical or health record.^1^ For prescribing of medication, CPOE can incorporate clinical decision support to improve medication and patient safety.^2–7^ Examples of clinical decision support to improve medication safety are standard order sentences, dosing calculators, drug interaction and allergy alerts, and evidence-based treatment recommendations.^8^ However, CPOE systems require ongoing optimisation to ensure those patient safety gains are maintained and improved.^9–11^ To achieve this, technology-related errors (TREs)^12–17^ need to be addressed as part of CPOE optimisation.^11, 18^

TREs, also termed system-related errors, technology-induced errors or computer-related errors, are errors that arise from “the use and functionality of [systems] which would be unlikely or unable to occur in paper-based medication ordering systems”.^16^ TREs can significantly curb the benefits of CPOE – they have been reported to account for between 1.2% and 77.7% of all medication errors,^5, 15, 16^ and can persist for many years after CPOE implementation.^17^ TREs may be an indication of system usability issues, as a result of CPOE not supporting users to complete tasks efficiently and effectively. Thus, addressing TREs could lead to substantial improvements in both patient safety and system usability.

To effectively address TREs, a clear understanding of their underlying mechanisms is required. For example, possible mechanisms for a wrong dose error of a transdermal fentanyl patch may be an incorrect selection from a drop-down menu or not changing a default setting for the ‘strength’ field of the order. Depending upon the mechanism, the strategy to prevent this TRE would differ. Evidence of selection errors, for example, can inform changes to drop-down menu arrangement^17, 19^ or the introduction of new system logic to minimise options displayed in a drop-down menu. Importantly, the underlying mechanisms of TREs are distinct from the manifestation of the TRE, e.g. a wrong dose or wrong drug error.^16^

Multiple approaches to classify TREs have been developed, predominantly using data from incident reporting systems.^20–24^ These classifications vary widely in their level of detail, grouping of categories, purpose and use. Many are intended for classifying issues with health IT systems more broadly and not specifically issues with the medication prescribing process.^21, 22, 25^ As a result, they typically do not contain sufficient detail to facilitate identification of areas for CPOE optimisation. Furthermore, previous TRE classifications conflate error types and mechanisms, reducing the utility of results.

Previously, we developed a dual classification for TREs using data on 1164 prescribing errors from two adult hospitals with different CPOE systems.^2, 16^ This classification system categorised errors in two dimensions, as shown in Figure 1: the error manifestation (e.g. wrong dose) and the error’s underlying mechanisms (e.g. selection error). The mechanism classification categories described ‘how’ errors occurred with the aim of allowing system designers to understand the specific CPOE features that are associated with TREs, and hence support the design of potential solutions. Applying the mechanism classification, for example, showed that CPOE designs with fewer drop-down menu options had a lower rate of incorrect selection of menu options.^16^

**Figure 1:**
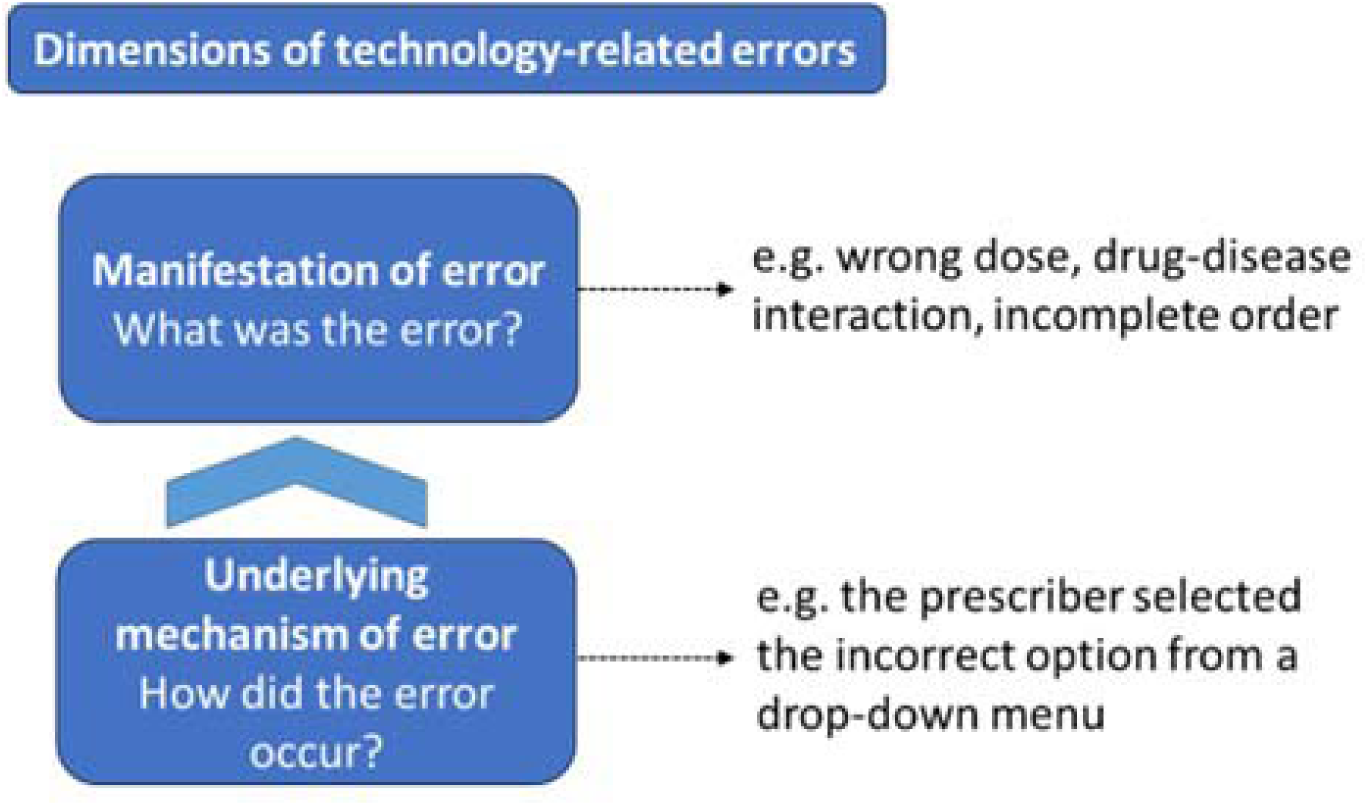
Two dimensions of technology-related errors with examples

The most frequently reported dimension of TREs, in studies of medication errors and in incident reports, is the manifestation of the error (Figure 1).^4, 5, 26^ This is also the most visible dimension of TREs in clinical practice and incident reports. How the error occurred in the CPOE, i.e. the underlying mechanism of the error, is a less visible dimension of TREs. Our classification of mechanisms of TREs brought to the fore information on how TREs occurred and allowed for a systematic examination of where the CPOE optimisation could focus.

Our original TRE mechanism classification, however, was developed almost a decade ago, and as CPOE systems have become more sophisticated, the tools used to evaluate the systems should also be reviewed and updated. The applicability of the mechanism classification to paediatrics also had never been tested. Building on previous work, our aim was to update our TRE mechanism classification, incorporating new data generated from a large paediatric dataset, and to assess the reliability with which reviewers could independently apply the classification.

## METHODS

### Data used to inform classification development

We conducted a secondary analysis of prescribing error data generated from a study at a paediatric referral hospital in Sydney, Australia. The hospital implemented a CPOE module onto an existing electronic medical record (Cerner) to support electronic prescribing and medication administration. Prescribing errors were identified by research pharmacists with clinical practice experience in a retrospective audit of medication orders during a 10-week CPOE implementation period in 2016, then again over a corresponding period in 2017, one year after the introduction of the CPOE.^7^ All prescribing errors in the dataset had an assigned clinical (e.g. wrong dose) or procedural (e.g. unclear order) error category and data recorded using a structured data collection form.^7^ Errors from the 2016 and 2017 datasets were used for classification development, with a sample of errors from 2017 used for inter-rater reliability testing of the updated classification.

### Classification development

In total 1696 prescribing errors were reviewed independently by two clinical pharmacist reviewers (AM and EF) to assess if they were technology-related. A TRE was defined as a prescribing error “where there was a high probability that the functionality or design of the [system] contributed to the error”,^16^ including errors arising from new work processes and changes in prescribing practices that were implemented with the introduction of the CPOE.^27^ Essentially, TREs were those errors unlikely or not possible to occur with paper-based prescribing.^17^ This definition focuses only on the functionality of the current CPOE and does not include errors that the system failed to prevent due to lack of decision support.^23^

The mechanisms of the technology-related prescribing errors were then categorised according to the previously developed classification, which consisted of four main mechanism categories, with 10 sub-categories.^16^ Assessment and classification of error mechanisms were performed by the two reviewers independently, in blocks of 50 errors. Mechanisms not captured in the existing classification were documented by reviewers. Following review of each block, comparison of results, discussion of the classification categories and identification of new categories and sub-categories took place. The reviewers continued to apply the classification, including new categories, to the dataset until no new mechanisms were identified. The reviewers also assessed whether more than one mechanism category could be applied to a single TRE when applying the classification.

In order to conduct the error assessments, the reviewers had access to the medication orders with errors in the CPOE (Cerner™), including details of how an order was entered (e.g. using order sentences or ad-hoc order entry; whether a dose calculator was used, etc), the order time and date, prescriber details and patient notes. Additionally, access to a testing environment of the CPOE system enabled simulated prescribing to support identification of TRE mechanisms, and particularly the testing of new mechanisms. Table 1 provides an overview of the how the two sources were used as part of a two-step process for assessing TREs and related mechanisms. Examples of investigational questions reviewers asked to guide the process are provided. Supplementary file 1 provides detailed worked examples using these steps.

**Table 1:**
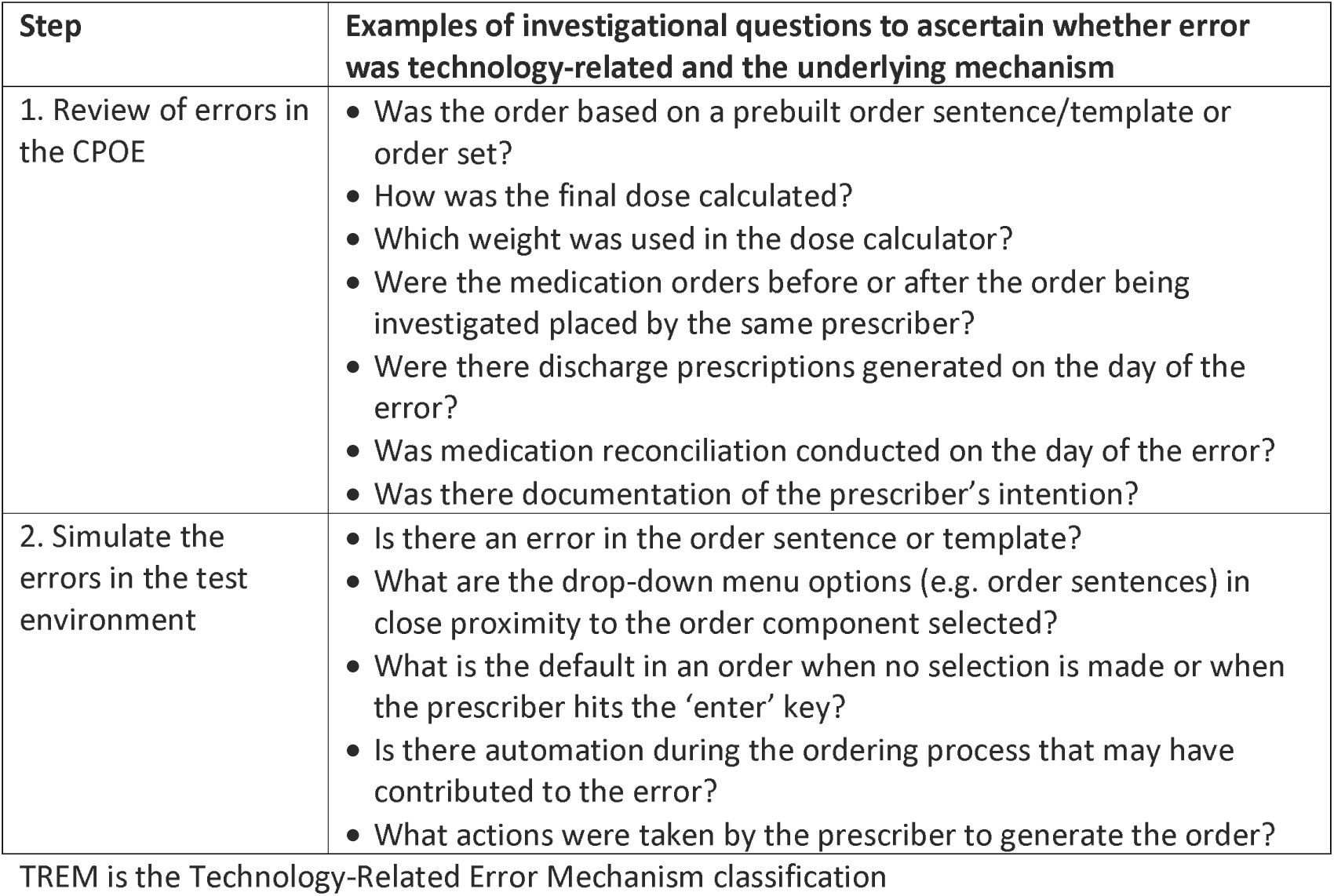
Two-step process and examples of investigational questions for applying the TREM classification.

The mechanism categories and sub-categories developed were reviewed by and discussed with the broader research team in an iterative manner during development. The final classification was presented to a project steering committee consisting of doctors (paediatricians, pharmacologists, and other medical specialists), nurses, pharmacists, patient safety and human factors experts, hospital executives and informaticians. Input from the committee ensured face validity of the mechanism categories, definitions and examples, and that they were easily understood by both clinicians and non-clinical members.

### Assessing the reliability of the application of the new TREM classification

Following the development of the final classification, we conducted inter-rater reliability testing to determine the consistency with which reviewers could independently apply the classification to a new dataset. A sample of 231 clinical prescribing errors from the 2017 data set was extracted and independently assessed by two reviewers. The errors identified as TREs had their underlying mechanisms classified using the updated classification. This sample comprised a wide range of error manifestations including, but not limited to, dose, route, frequency, timing and duplication errors. Cohen’s kappa scores were calculated for: 1) whether an error was technology-related (Yes/No), and; 2) the TRE mechanism categories assigned.

## RESULTS

### The Technology-Related prescribing Error Mechanism (TREM) classification

The original mechanism classification comprised four major categories and 10 sub-categories.^16^ Through the review of paediatric prescribing errors, our classification was extended to seven major mechanism categories with 19 sub-categories. Table 2 provides an overview of the Technology-Related Error Mechanism (TREM) classification, and a more detailed description of mechanism sub-categories with examples is provided in Supplementary file 2.

**Table 2:**
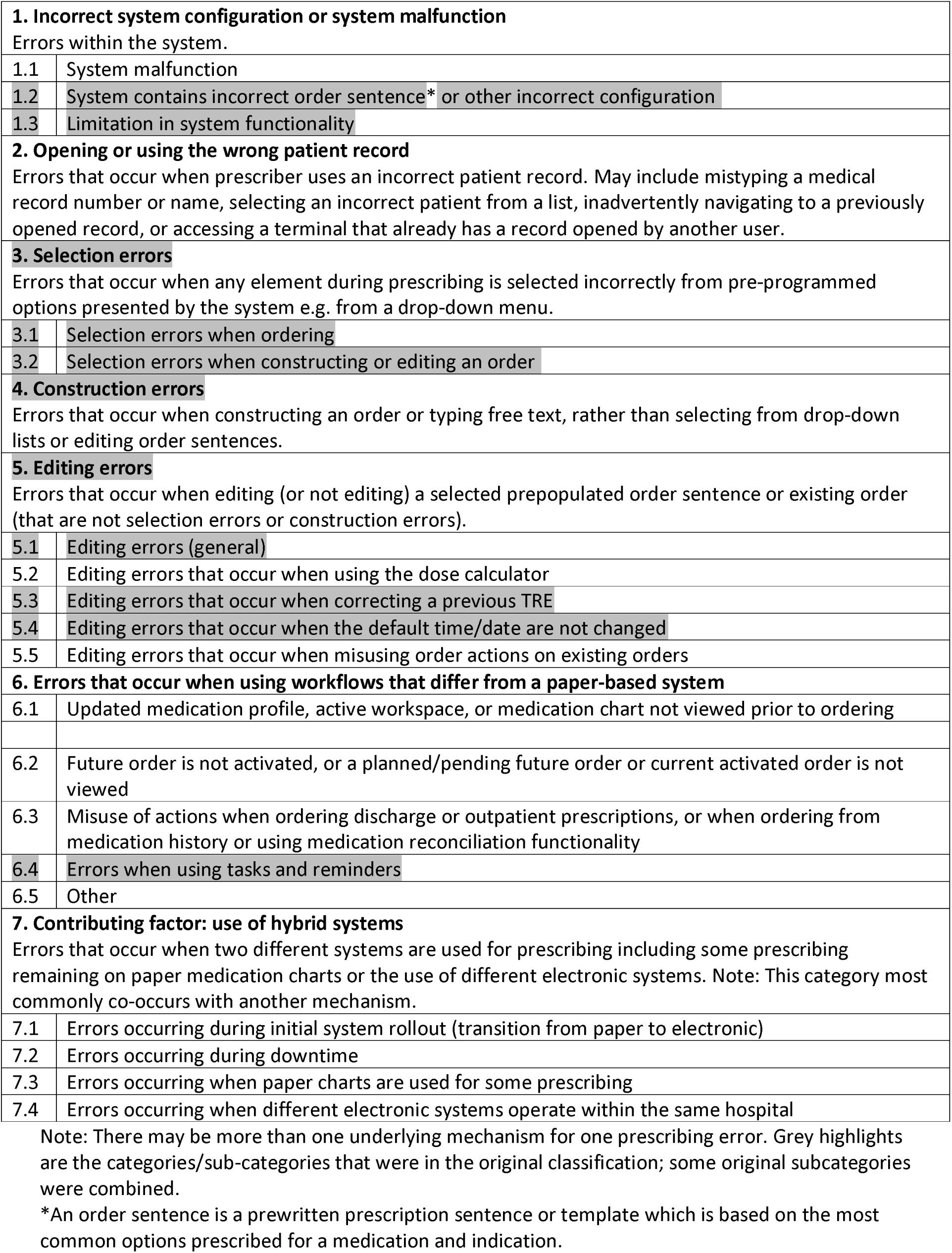
Major categories and sub-categories of the Technology-Related Error Mechanism (TREM) classification.

|  |  |
| --- | --- |
| **1. Incorrect system configuration or system malfunction** |  |
| Errors within the system. |  |
| 1.1 | System malfunction |
| 1.2 | System contains incorrect order sentence* or other incorrect configuration |
| 1.3 | Limitation in system functionality |
| **2. Opening or using the wrong patient record** |  |
| Errors that occur when prescriber uses an incorrect patient record. May include mistyping a medical record number or name, selecting an incorrect patient from a list, inadvertently navigating to a previously opened record, or accessing a terminal that already has a record opened by another user. |  |
| **3. Selection errors** |  |
| Errors that occur when any element during prescribing is selected incorrectly from pre-programmed options presented by the system e.g. from a drop-down menu. |  |
| 3.1 | Selection errors when ordering |
| 3.2 | Selection errors when constructing or editing an order |
| **4. Construction errors** |  |
| Errors that occur when constructing an order or typing free text, rather than selecting from drop-down lists or editing order sentences. |  |
| **5. Editing errors** |  |
| Errors that occur when editing (or not editing) a selected prepopulated order sentence or existing order (that are not selection errors or construction errors). |  |
| 5.1 | Editing errors (general) |
| 5.2 | Editing errors that occur when using the dose calculator |
| 5.3 | Editing errors that occur when correcting a previous TRE |
| 5.4 | Editing errors that occur when the default time/date are not changed |
| 5.5 | Editing errors that occur when misusing order actions on existing orders |
| **6. Errors that occur when using workflows that differ from a paper-based system** |  |
| 6.1 | Updated medication profile, active workspace, or medication chart not viewed prior to ordering |
| 6.2 | Future order is not activated, or a planned/pending future order or current activated order is not viewed |
| 6.3 | Misuse of actions when ordering discharge or outpatient prescriptions, or when ordering from medication history or using medication reconciliation functionality |
| 6.4 | Errors when using tasks and reminders |
| 6.5 | Other |
| **7. Contributing factor: use of hybrid systems** |  |
| Errors that occur when two different systems are used for prescribing including some prescribing remaining on paper medication charts or the use of different electronic systems. Note: This category most commonly co-occurs with another mechanism. |  |
| 7.1 | Errors occurring during initial system rollout (transition from paper to electronic) |
| 7.2 | Errors occurring during downtime |
| 7.3 | Errors occurring when paper charts are used for some prescribing |
| 7.4 | Errors occurring when different electronic systems operate within the same hospital |

Note: There may be more than one underlying mechanism for one prescribing error. Grey highlights are the categories/sub-categories that were in the original classification; some original subcategories were combined.
\*An order sentence is a prewritten prescription sentence or template which is based on the most common options prescribed for a medication and indication.

### Significant updates to the mechanism classification

Four categories were added to the mechanism classification, and sub-categories were reorganised and expanded (the categories and/or sub-categories from the original classification are shaded in grey in Table 2). Firstly, we created a new category describing ‘Incorrect system configuration or system malfunction’ that encompassed two sub-categories of the previous classification. Secondly, category two, ‘Opening or using the wrong patient record’ acknowledges that wrong patient errors are more likely with CPOE than on paper medication charts. This mechanism includes TREs that may have occurred with a number of processes, however, with retrospective record review, it is difficult to identify exactly which process led to an individual error. For example, a prescriber may have inadvertently ordered medication on a patient record already open in the CPOE or selected the wrong patient record. Thirdly, category six, ‘Errors that occur when using workflows that differ from a paper-based system’, was refined and encompasses a set of mechanisms where staff misused CPOE features that are new workflows compared to paper-based prescribing. For example, these errors included those when attempting to create discharge prescriptions or enter reminders on the chart. Lastly, category seven, ‘Contributing factor: use of hybrid systems’, was added recognising that in some jurisdictions, hospitals may use paper prescribing for complex medications (e.g. insulin sliding scale orders) in addition to CPOE or employ different CPOE systems in different units (e.g. in intensive care versus general wards; emergency department versus general wards). Errors relating to hybrid systems often co-occur with other mechanism categories, as hybrid systems are a contributing factor to TREs.

The application of more than one mechanism for each error was also a new development for the expanded classification. Results of our testing indicated that in some situations more than one underlying mechanism may occur simultaneously or sequentially. Of all TREs identified over both time periods (n=526), 24.1% (n=127) involved more than one mechanism. By way of example, a duplicate drug therapy error may have an underlying mechanism of ‘Contributing factor: use of hybrid systems’, as well as ‘Errors when using tasks and reminders’ when there was incorrect use of a ‘placeholder’ in the CPOE system to alert users that a paper order existed. Similarly, a dose error involving incorrect selection from a drop-down menu could also be associated with incorrect editing within the dose calculator.

### Inter-rater reliability in the application of the TREM classification

There was moderate to strong agreement^28^ between the two clinical pharmacist reviewers when they independently assessed a sub-sample of 231 errors from the 2017 dataset to determine whether an error was a TRE or not, with a kappa score of 0.79 (95% CI: 0.71-0.88).

A total of 63 prescribing errors were identified by both reviewers as TREs. The TRE mechanisms of these 63 cases were then also independently reviewed and assigned one or more mechanism categories. For the mechanism categories, the kappa score was 0.80 (95% CI: 0.68-0.92), indicating moderate to strong agreement.^28^

Disagreements for assessing whether an error was a TRE and assigning a mechanism often revealed that a more detailed review of the medical record was required to understand how an error occurred. For example, a dose error due to an incorrect weight being entered was interpreted by one reviewer as a TRE (construction error); however, a more detailed examination of the record uncovered that the weight used was accurately transcribed from the emergency department notes but was out of date, and thus was not a technology-related error. Disagreements with assigning a mechanism category most frequently occurred when multiple mechanisms were assigned. For example, an assignment of a hybrid system category often co-occurred with misuse of placeholders; however, in a small number of instances the placeholder was correctly applied in the CPOE, but a duplicate error occurred as there was a paper chart and CPOE in use. Again, the initial disagreement between reviewers required a more careful examination of the medical record.

## DISCUSSION

The unintended consequences of health IT, including TREs, have been recognised for over two decades.^12, 13^ Addressing TREs is a key component of CPOE optimisation.^11, 18^ The TREM classification reported in this paper provides a systematic means to understand how technology-related prescribing errors occur, allowing targeted CPOE modifications to be made. The classification was initially developed through a review of 1164 prescribing errors at two adult hospitals using two different CPOE systems.^16^ It has now been expanded with a review of prescribing errors at a tertiary paediatric hospital.

Updating the classification with the prescribing error data from a paediatric hospital resulted in 3 new mechanism categories and re-organisation of sub-categories. One new sub-category was specific to paediatric CPOE functionality, ‘Editing errors that occur when using the dose calculator’, with dose calculators used for the vast majority of paediatric prescribing. The other updates to the classification could equally apply to the adult setting, however we have also added examples from paediatrics. For instance, we encountered construction errors where the weight of the child was entered incorrectly when constructing an order. Similarly, we found that off-label prescribing, i.e. use of medications for indications for which they are not licensed, was more frequent among the paediatric population requiring editing of pre-programmed order sentences or constructing orders. However, the core elements of the classification, first applied to CPOE systems in two adult hospitals, remained applicable to the paediatric CPOE when applied eight years later, providing an indication of its utility and relevance despite design changes to CPOE systems during that time. Our updated classification is more comprehensive as a result of including a contemporary dataset of prescribing errors from another setting and can be applied to CPOE for both paediatric and adult populations.

The TREM classification focuses on ‘how’ TREs occur, which allows for targeted CPOE modifications. We have demonstrated the feasibility of informing CPOE modifications using the TREM classification. Recommendations arising from our work have already been implemented within the study hospital’s CPOE; for example, changing the drop-down menu options for the intravenous route, and new design options to avoid inadvertent selection of controlled release opioids. Multiple optimisation strategies informed by the TRE data have been formulated and published online in the Health Innovation Series to allow greater dissemination of specific system optimisation recommendations and user tips.^29–32^ Data on TRE mechanisms would also be a powerful addition to human factors approaches for evaluating and analysing CPOE systems, and in the design of solutions to address TREs.^33^

Research shows that health IT managers are flooded with requests to modify health IT systems, but often have limited resources to respond to these requests.^9–11, 34^ Thus, prioritisation of CPOE modifications based upon factors such as the safety, efficiency and frequency of issues identified is required. It is also recognised that data to support the identification, prioritisation and evaluation of CPOE optimisation to address TREs are needed. To date, the majority of data on TREs has originated from voluntary incident reports from hospitals or regulatory bodies, such as the Food and Drug Administration.^18^ While incident reports can provide valuable insights into TREs, they cannot provide information on the relative frequency with which TREs are occurring or allow for meaningful comparisons between organisations.^23, 35, 36^ Incident reporting systems are also less likely to capture TREs with minimal or no patient impact. However, these less serious TREs may significantly affect health IT usability, and thus should be addressed to improve user experience by removing obstacles to task completion and improving workflows.

In order to effectively prioritise CPOE optimisation activities, representative data on multiple dimensions of TREs are required. In Figure 1, we showed two key dimensions of TREs.

Figure 2 expands on this concept with a third dimension of TREs, i.e. the outcomes of the error, and also shows other examples of the error manifestation. In work spanning the last two decades, our team has developed classifications that can be used to describe TRE dimensions, and examples of these classifications are provided in Figure 2.^2–6, 16, 26, 36–38^

**Figure 2:**
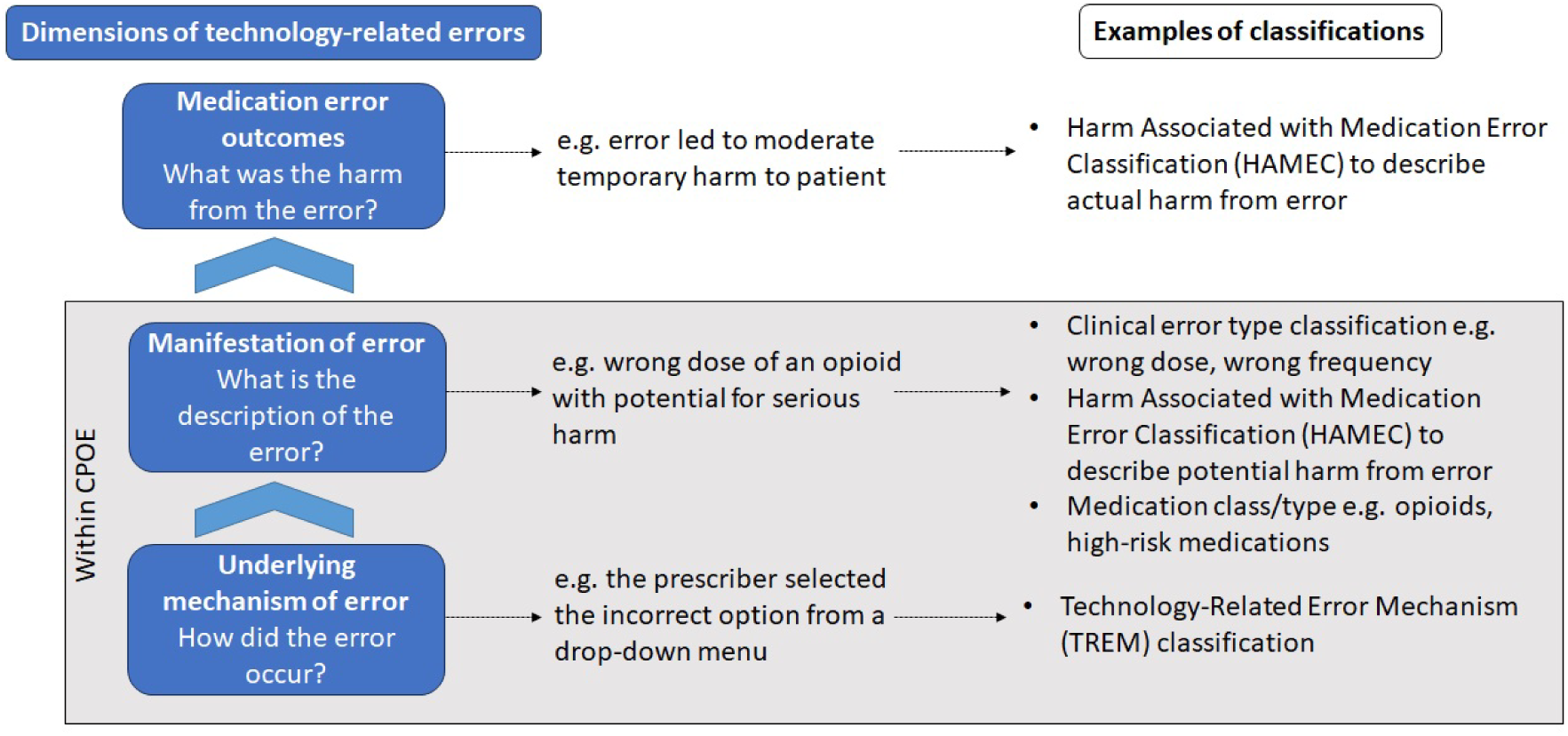
Dimensions of technology-related errors with descriptions and classification examples

Armed with data on multiple dimensions of TREs, CPOE managers could prioritise optimisation activities according to their goals or known areas of risk. For example, they could target the most frequent clinical error type by examining the underlying mechanism. Similarly, to reduce TREs with high-risk medication orders, such as high strength potassium fluids or opioids, the underlying mechanisms of these errors could inform CPOE optimisation to support prescribers when ordering. Table 3 shows further examples of how CPOE optimisation goals can be mapped to the TRE dimensions shown in Figure 2.

**Table 3:**
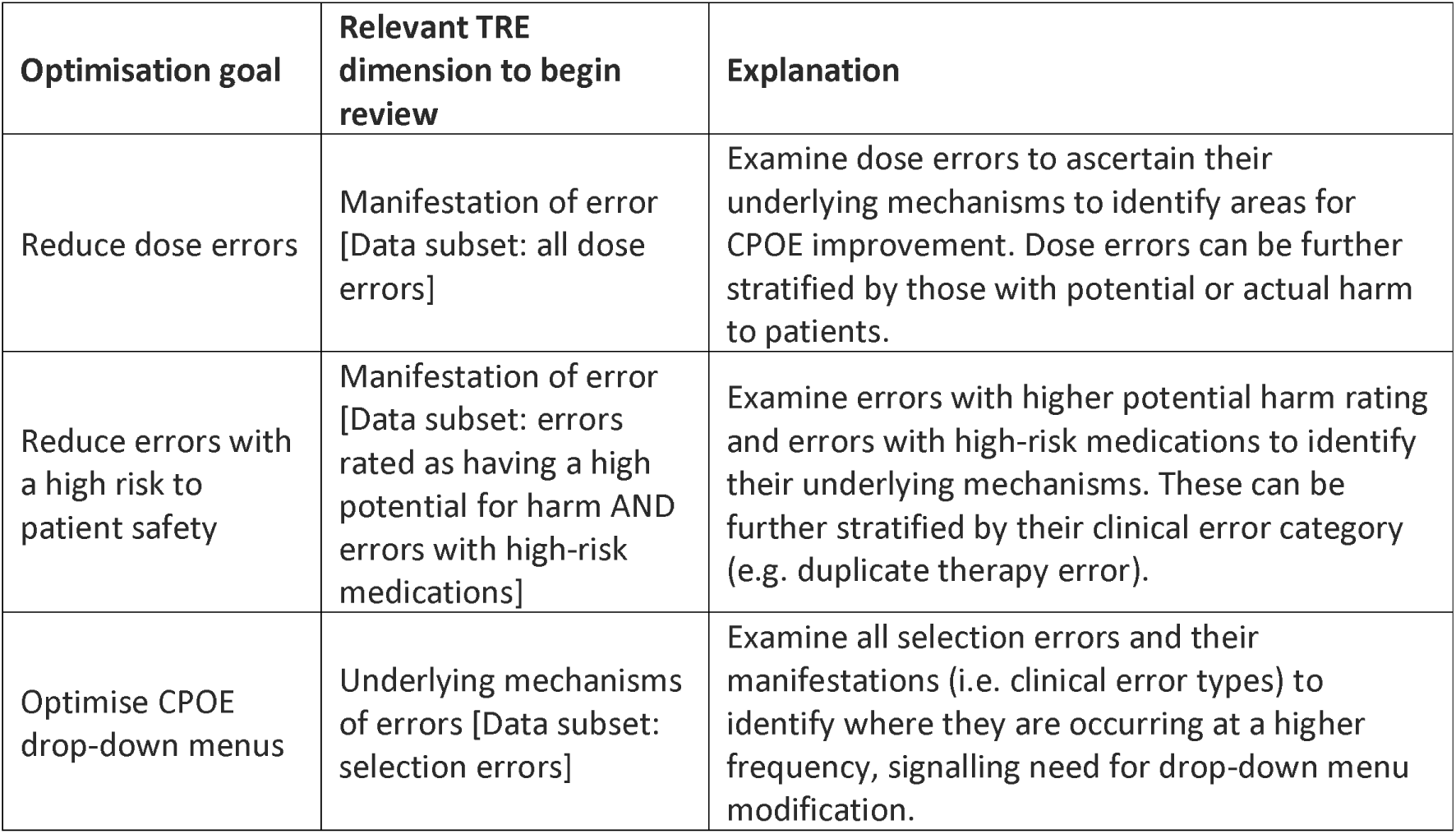
Examples of how dimensions of technology-related prescribing errors can be used to prioritise CPOE optimisation goals.

| Optimisation goal | Relevant TRE dimension to begin review | Explanation |
| --- | --- | --- |
| Reduce dose errors | Manifestation of error [Data subset: all dose errors] | Examine dose errors to ascertain their underlying mechanisms to identify areas for CPOE improvement. Dose errors can be further stratified by those with potential or actual harm to patients. |
| Reduce errors with a high risk to patient safety | Manifestation of error [Data subset: errors rated as having a high potential for harm AND errors with high-risk medications] | Examine errors with higher potential harm rating and errors with high-risk medications to identify their underlying mechanisms. These can be further stratified by their clinical error category (e.g. duplicate therapy error). |
| Optimise CPOE drop-down menus | Underlying mechanisms of errors [Data subset: selection errors] | Examine all selection errors and their manifestations (i.e. clinical error types) to identify where they are occurring at a higher frequency, signalling need for drop-down menu modification. |

To apply the TREM classification, data on prescribing errors within a CPOE are required to identify and classify the multiple dimensions of TREs. It is important, however, to recognise that this may be a labour-intensive process and thus, potential users of the classification should consider how best to capture error data within existing medication safety processes e.g. pharmacist review of medication orders. Future work could explore automation of TRE detection to reduce record review workload for potential users. Alternately, the TREM classification could also be used to proactively improve ordering for high-risk medications and prescribing scenarios. For example, the risk of selection errors could be examined for high-risk medications and changes made to limit options in drop-down menus.^31^

### Strengths and limitations

The TREM classification’s strengths are that it has been developed based on empirical evidence from both adult and paediatric inpatient populations, across a variety of hospital wards using two commercial CPOE systems. However, there are some limitations. The classification was developed using inpatient orders only, and applicability for the analysis of discharge and outpatient orders is yet to be determined. Though incident reports provide a readily available source of data on medication errors for most institutions, whether the classification can be applied to errors reported in incident reports remains to be tested.

Prescribing error data from chart review provides a very detailed classification of error types, while incident reports often have limited information, and reporters may lack understanding or ability to fully describe how the errors occurred in the CPOE.^23, 39^ We also acknowledge that despite being informed by data from multiple settings, there may be other mechanism categories not captured by the classification, for example due to the age of the data used, methods being restricted to retrospective record review or differing functionality in other CPOE systems.^40^ However, the assessment of paediatric prescribing errors yielded further categories of TRE mechanisms compared with those generated using prescribing errors in adult hospitals, demonstrating the importance of considering the other contexts when examining classifications of medication errors, particularly for workflows that may differ to those when prescribing for adults. Lastly, inter-rater reliability was conducted between two pharmacist researchers who were involved in updating the classification. An assessment of interrater reliability with people external to the project would be a useful next step.

## Conclusion

The need to address TREs has been recognised for over two decades and more recently as a key consideration for CPOE optimisation. We reviewed and updated a TRE mechanism classification by incorporating new data generated from a large paediatric dataset, and demonstrated its reliability and applicability to the paediatric setting. Our classification of the underlying mechanisms of TREs facilitates an understanding of ‘how’ errors occurred, thus identifying where system design requires modification. When used with information about other dimensions of TREs, it allows the prioritisation of optimisation goals targeting TREs. We envisage that the application of the TREM classification as part of an ongoing technology audit will identify CPOE issues and allow evaluation of CPOE modifications.

## Data Availability

All data produced in the present study are available upon reasonable request to the authors.

## ACKNOWLEDGEMENTS

MZR and AM contributed equally to this manuscript and are joint first authors.

## AUTHOR CONTRIBUTIONS

JIW, MTB, LL, MZR conceptualized the study. AM and EF conducted error assessment and review. MZR, MTB, AM, EF and PG contributed to classification development. LL analysed data to assess inter-rater reliability of the application of the classification. MZR and AM presented the classification to project steering group. AM and MZR jointly drafted the manuscript and thus are equal first authors. All authors contributed to the manuscript drafting and revision, and approved the final version of the manuscript.

