## Supplementary file 1 for "Understanding technology-related prescribing errors for system optimisation: the Technology-Related Error Mechanism (TREM) classification"

### SUPPLEMENTARY FILE 1: Guide to applying the Technology-Related Error Mechanism (TREM) classification

**Resources required to apply the TREM classification**

To determine technology-related prescribing errors and apply the Technology-Related Error Mechanism (TREM) classification, investigators should have access to the following:

- Details of prescribing errors, including the error category (e.g. wrong dose)
- A rating of potential and/or actual harm for each error is beneficial, but not required to assess whether errors are technology-related and the underlying mechanisms of TREs
- Access to the computerised provider order entry (CPOE) records to view the orders with errors
- Access to a testing environment of the CPOE system

**Steps to be taken to apply the TREM classification**

The steps taken to determine whether an error is technology-related and assign an underlying mechanism are outlined in Table S2, along with examples of investigational questions.

**Table S2: Two-step process and examples of investigational questions for applying the Technology-Related Error Mechanism (TREM) classification**

| Step | Examples of investigational questions to ascertain whether an error was technology-related and the underlying mechanism |
|---|---|
| 1. Review of error in the CPOE | • Was the order based on a prebuilt order sentence/template or order set?<br>• How was the final dose calculated?<br>• Which weight was used in the dose calculator?<br>• Were the medication orders before or after the order being investigated placed by the same prescriber?<br>• Were there discharge prescriptions generated on the day of the error?<br>• Was medication reconciliation conducted on the day of the error?<br>• Was there documentation of the prescriber's intention? |
| 2. Simulate the error in the test environment | • Is there an error in the order sentence or template?<br>• What are the drop-down menu options (e.g. order sentences) in close proximity to the order component selected?<br>• What is the default in an order when no selection is made or when the prescriber hits the 'enter' key?<br>• Is there automation during the ordering process that may have contributed to the error?<br>• What actions were taken by the prescriber to generate the order? |

The following pages provide worked examples of error investigations to guide users in the application of the TREM classification:

- Underlying mechanism example 1: System contains an incorrect order sentence or other incorrect configuration
- Underlying mechanism example 2: Construction error
- Underlying mechanism example 3: Editing errors relating to the dose calculator
- Underlying mechanism example 4: Selection error
- Example 5: Error not classified as a TRE

#### Underlying mechanism example 1: System contains an incorrect order sentence or other incorrect configuration

**Clinical error type:** Wrong dose

<u>Details</u>: *fentanyl 72 microg, intranasal-both, ONCE*, indicating one spray of 72 microg per nostril giving a total dose of 144 microg. However, this should be one spray of 72 microg in ONE nostril.

**fentanyl (fentanyl nasal spray)**
72 microg, Intranasal-Both, ONCE, Stop Date: 03-Jul-16 14:20:00, STAT, Moderate-Severe Pain, Use a 600 microg/2mL intranasal vial to make up the fentanyl nasal spray as per CHW ED guideline. | 72 microg @1429

##### STEP 1: REVIEW ORDER IN THE CPOE

**Investigational question 1: Was the order based on a prebuilt order sentence?**

On investigation it was noted that the order was based on a prebuilt order sentence, as indicated by the inclusion of information such as the 'Drug Dosage Guidelines' and 'Dose Limits' embedded in the order sentence. These components were usually not completed by a prescriber when constructing an order without an order sentence.

**Details**

| | |
|---|---|
| Strength Dose | 72 |
| Strength Dose Unit | microg |
| Target dose | 1.5 microg/kg |
| Actual dose | 1.5 microg/kg |
| Route of Administration | Intranasal-Both |
| Drug Form | Spray |
| Frequency | ONCE |
| First Dose URGENCY | STAT |
| FIRST DOSE Date/Time | 03/07/2016 14:20 AEST |
| Stop Date/Time | 03/07/2016 14:20 AEST |
| Indication | Moderate-Severe Pain |
| Special Instructions | Use a 600 microg/2mL intranasal vial to make up the fentanyl nasal spray as per CHW ED guideline. |
| Drug Dosage Guidelines | CHW ED Intranasal Fentanyl |
| Dose Limit | 100 micrograms |

##### STEP 2: SIMULATE THE ERROR IN THE TEST ENVIRONMENT

**Investigational question: Is there an error in the order sentence or template?**

On examination of the test environment and the available order sentences it was apparent that the order sentence had been prebuilt with the route 'intranasal-both'.

##### CONCLUSIONS

The underlying mechanism of the error was '1.2 System contains incorrect order sentence or other incorrect configuration'.

#### Underlying mechanism example 2: Construction error

**Clinical error type:** Wrong dose

<u>Details</u>: *Oxycodone 0.6 gm, Oral/Nasogastric, 6 hourly* prescribed resulting in a 1000 times overdose as the units should have been ‘mg’ (and not ‘gm’).

| | |
|---|---|
| oxycodone<br>0.6gm, Oral / Nasogastric, 6 hourly, analgesia | 0.6 mg @1200 |
| | Pain Score: 3 |
| | Not Done: Not appropriate at this time @1900 |

**STEP 1: REVIEW ORDER IN THE CPOE**

**Investigational question 1: Was the order based on a prebuilt order sentence?**

On investigation it was noted that the order was <u>not</u> based on a prebuilt order sentence due to the absence of the usual order sentence information such as ‘Drug Dosage Guidelines’ and ‘Dose Limits’. Therefore, the prescriber created the order using the ‘free-text’ prescribing option, eliminating mechanism categories related to order sentences e.g. Selection errors.

**Details**

| | |
|---|---|
| Freetext Dose | 0.6gm |
| Route of Administration | Oral / Nasogastric |
| Frequency | 6 hourly |
| First Dose URGENCY | Routine |
| FIRST DOSE Date/Time | 03/07/2016 13:00 AEST |
| Indication | analgesia |
| Instructions Replace Required Details | No |
| Next dose date and time | 03/07/2016 13:00 AEST |

**Investigational question 2: Was there information in the order as to how the dose had been determined?**

In this case the prescriber did not use the dose calculator to calculate the final dose, as there was no 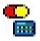 symbol next to the dose and the dose is labelled ‘Freetext Dose’. Furthermore, the dose calculator is not presented to the prescriber when creating free text orders.

**CONCLUSIONS**

The underlying mechanism of the error was ‘4. Construction errors’.

#### Underlying mechanism example 3: Editing errors that occur when using the dose calculator

**Clinical error type**: Wrong dose

Details: *metronidazole IV intermittent 810 mg 12 hourly* prescribed for a 14-year old (weight: 64.6 kg, height: 174 cm). The dose limit for IV metronidazole is 500 mg per dose according to guidelines.

**metronidazole**
810 mg, IV Intermittent Infusion, 12 hourly, Order Duration: 24 hr(s), Wound infection, Surgical Prophylaxis, Stop Date: 27-Aug-20 15:59:00

**STEP 1: REVIEW ORDER IN THE CPOE**

**Investigational question 1: Was the order based on a prebuilt order sentence?**

On investigation it was noted that the order was based on a prebuilt order sentence, as indicated by the inclusion of information such as the ‘Drug Dosage Guidelines’ and ‘Dose Limits’ embedded in the order sentence. These components were usually not completed by a prescriber when constructing an order without an order sentence.

| | |
|---|---|
| Strength Dose | 810 |
| Strength Dose Unit | mg |
| Route of Administration | IV Intermittent Infusion |
| Frequency | 12 hourly |
| First Dose URGENCY | Routine |
| FIRST DOSE Date/Time | 26/08/2020 16:00 AEST |
| PRN | No |
| Duration | 24 |
| Duration Unit | hr(s) |
| Stop Date/Time | 27/08/2020 15:59 AEST |
| AMS Indication | Wound infection |
| Indication | Surgical Prophylaxis |
| AMS Approval | intraoperative |
| Use Patient's Own Medication | No |
| Order for Future Visit | No |
| Drug Dosage Guidelines | M4K: 12.5mg/kg/dose q12h |
| Dose Limit | 500mg per Dose |

In addition it was noted that the order sentence was part of a PowerPlan (as indicated by the following symbol attached to the order) , and by the pathway description in the 'Additional Info' tab attached to the order:

Comments | History | Validation | Details | Ingredients | **Additional Info** | Results

| | |
|---|---|
| Ordered As | metronidazole |
| Start Date/Time | 26/08/2020 16:00 AEST |
| Stop Date/Time | 26/08/2020 15:09 AEST |
| Frequency | 12 hourly |
| Order ID | 695983851 |

**Pathways**

| | |
|---|---|
| Post-operative Inpatient Orders [redacted] | 231204629 |

**Investigational question 2: Was there information in the order as to how the dose had been determined?**

In this case the prescriber had used the dose calculator to calculate the final dose (as indicated by the symbol next to the dose). The dose calculator provided information on the target dose, the weight used to calculate the dose (in this case actual body weight) as well as the pre-programmed standard dose limit of 500 mg.

**metronidazole**

Dose Values

| | | | | |
|---|---|---|---|---|
| 1) Target dose: | 12.5 | mg/kg | | |
| 2) Calculated dose: | 807.5 | mg | | |
| 3) Dose Adjustment: | 807.5 | mg | 100 | % |
| 4) Final dose: | 810 | mg | 12.5387 | mg/kg |
| 5) Standard dose: | 500 | mg | 7.7399 | mg/kg |
| 6) Rounding rule: | Nearest ten | | | |
| 7) Adjust Reason: | | | | |
| 8) Route: | IV Intermittent Infusion | | | |

Order Comments

Reference Data

| | | | | |
|---|---|---|---|---|
| Date of birth: | 04/03/2006 | (14 Years) | | |
| Sex: | Male | | | |
| Race | Neither | | | |
| Height: | 174.9 | cm | Source: | 26/08/2020 11:23 174.90 cm Height |
| Actual weight: | 64.6 | kg | Source: | 26/08/2020 15:07 64.600 kg Weight Dosing |
| Adjusted weight: | 64.6 | kg | Adjustment: | Actual (no adjustment) |
| Serum creatinine: | 0 | umol/L | Source: | Manually entered |
| CrCl (est.): | | | Algorithm: | Schwartz - full term Missing data |
| | | | Weight Used for CrCl: | |
| Body surface area: | 1.77 | m2 | Algorithm: | Mosteller |

#### STEP 2: SIMULATE THE ERROR IN THE TEST ENVIRONMENT

**Investigational question: Is there an error in the order sentence or template?**

In this scenario, there was no error in the build or programming of the order sentence. When prescribing using the same order sentence selected by the prescriber and entering the child's weight and height, there was also no error in the dose calculator build.

**Investigational question: What are the drop-down menu options (e.g. order sentences) in close proximity to the order component selected?**

On examination of the test environment, it was noted that there were two order sentences for metronidazole IV to choose from depending on the child's weight (i.e. for a child weighing less than 40 kg and for a child greater than or equal to 40 kg). However, it was also noted that the order sentences are filtered based on the weight of the child recorded in the record, and thus only one order sentences would have been available to the prescriber, removing the likelihood of a selection error.

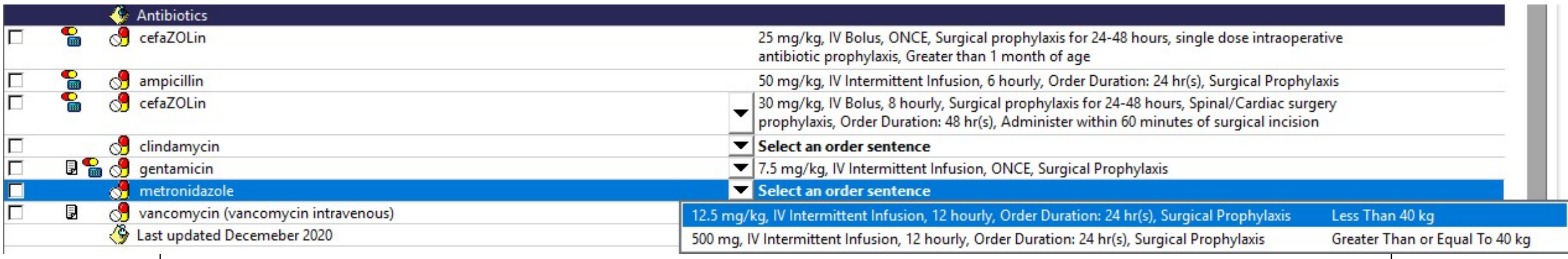


**Investigational question: What actions by the prescriber to generate the final dose in the order?**

It was determined that the dosage calculator was programmed with the standard dose limit of 500 mg (the 'standard dose'). However, the prescriber had to accept the dose limit by clicking the 'Apply Standard Dose' button. In this scenario, the prescriber instead clicked the 'Apply Dose' button, resulting in the final dose of 810 mg being applied to the order, instead of the intended 500 mg dose.

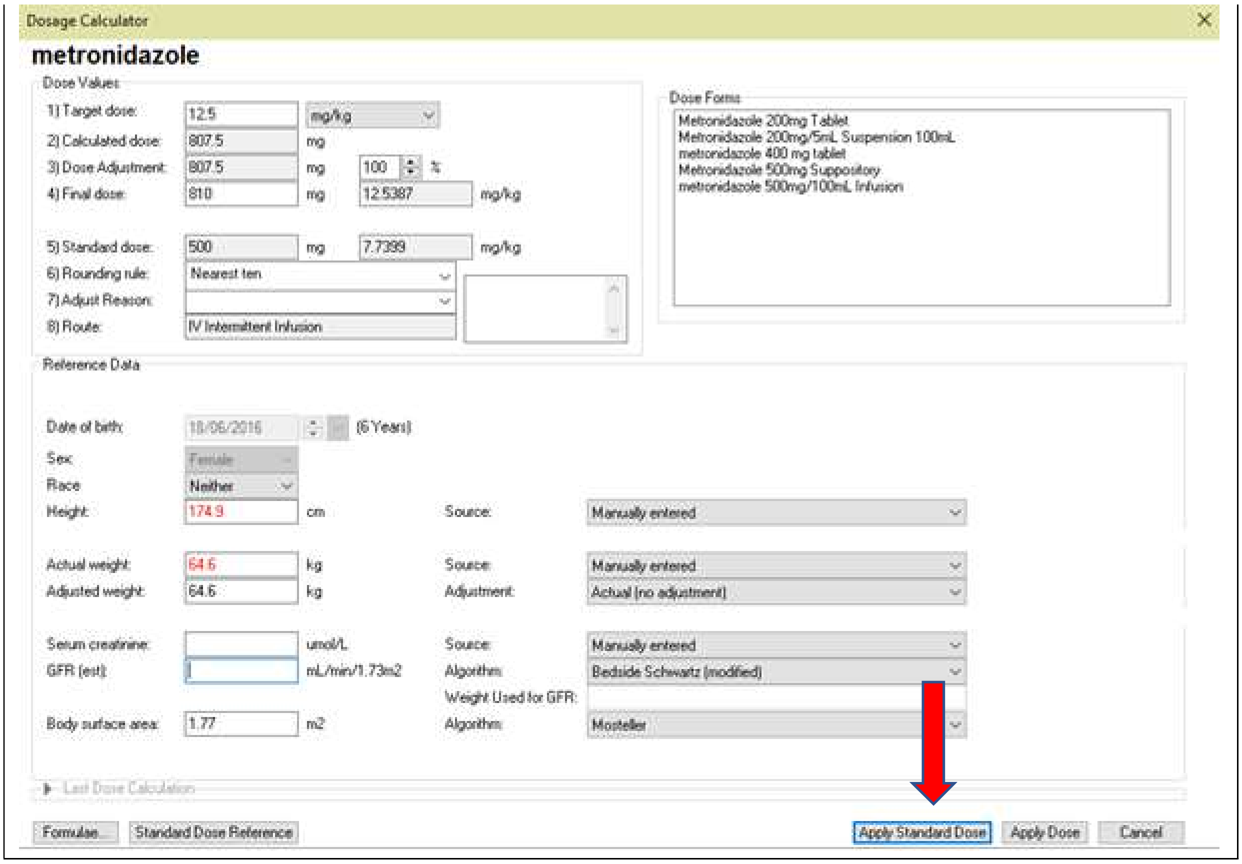


#### CONCLUSIONS

The underlying mechanism of the error was '5.2 Editing errors that occur when using the dose calculator'.

### Example 4: Selection error

**Clinical error type**: Wrong drug other

Details: *gentamicin inhalation 80 mg BD twice daily indication: cystic fibrosis*. The patient was being treated with intravenous therapy for a urinary tract infection, and did not have cystic fibrosis or require nebulised gentamicin.

gentamicin
80 mg, Nebulised Inhalation, BD, Cystic fibrosis

**STEP 1: REVIEW ORDER IN THE CPOE**

**Investigational question 1: Was the order based on a prebuilt order sentence?**

On investigation it was noted that the order was based on a prebuilt order sentence, as indicated by the inclusion of information such as the 'Drug Dosage Guidelines' embedded in the order sentence. These components were usually not completed by a prescriber when constructing an order without an order sentence.

**Details**

| | |
|---|---|
| Strength Dose | 80 |
| Strength Dose Unit | mg |
| Route of Administration | Nebulised Inhalation |
| Drug Form | Ampoule |
| Frequency | BD |
| First Dose URGENCY | Routine |
| FIRST DOSE Date/Time | 09/06/2016 20:00 AEST |
| Indication | Cystic fibrosis |
| Drug Dosage Guidelines | CHW: 80mg/dose nebulised q12h |
| Next dose date and time | 09/06/2016 20:00 AEST |

**Investigational question 1: What other medications were prescribed before and after the medication in error?**

On investigation it was noted that a second IV medication for urinary tract infection was prescribed at the same time as the order in error. In addition, the incorrect order had been ceased and reordered correctly as 80 mg IV intermittent infusion.

Scheduled

ampicillin
530 mg, IV Intermittent Infusion, 6 hourly, UTI

gentamicin
80 mg, Nebulised Inhalation, BD, Cystic fibrosis

gentamicin
80 mg, IV Intermittent Infusion, 24 hourly, UTI

#### STEP 2: SIMULATE THE ERROR IN THE TEST ENVIRONMENT

**Investigational question: What are the drop-down menu options (e.g. order sentences) relating to the order selected?**

On investigation it was noted that there were a number of gentamicin options presented to the prescriber to pick from in the drop-down menu. A prescriber intending to prescribe an 80 mg dose may inadvertently see and select the 80 mg nebulised dose rather than a mg/kg order sentence for IV therapy.

gentamicin
gentamicin 7.5 mg/kg, IV Intermittent Infusion, 24 hourly, Therapeutic drug monitoring is required [Greater Than or Equal To 1 month(s) And Less Than 43 kg]
gentamicin 320 mg, IV Intermittent Infusion, 24 hourly, Therapeutic drug monitoring is required [Greater Than or Equal To 1 month(s) And Greater Than or Equal To 43 kg]
gentamicin 2 mg/kg, IV Intermittent Infusion, ONCE, Surgical Prophylaxis, Administer within 2 hours before incision [Greater Than or Equal To 1 month(s)]
gentamicin 5 mg/kg, IV Intermittent Infusion, ONCE, Surgical Prophylaxis for Cardiac/Vascular/Lower Limb Surgery, Administer within 2 hours before incision [Greater Than or Equal To 1 month(s)]
gentamicin 7.5 mg/kg, IV Intermittent Infusion, 24 hourly, Febrile Neutropenia Oncology/BMT, Infuse over 5mins. Dose based on lean body weight for obese patients. [Greater Than or Equal To 1 month(s) And Less Than 42 kg]
gentamicin 320 mg, IV Intermittent Infusion, 24 hourly, Febrile Neutropenia Oncology/BMT, Infuse over 5mins. Dose based on lean body weight for obese patients. [Greater Than or Equal To 1 month(s) And Greater Than or Equal To 42 ...
gentamicin 10 mg/kg, IV Intermittent Infusion, 24 hourly, Cystic Fibrosis - Pseudomonal (acute) [Greater Than or Equal To 1 month(s)]
gentamicin 80 mg, Nebulised Inhalation, BD, Cystic fibrosis and non-CF bronchiectasis [Less Than 10 year(s)]
gentamicin 0.3% eye drops
gentamicin 0.3% eye drops 1 drop(s), Eye-Both, QID, Prophylaxis post superficial eye trauma or surgery
gentamicin 0.3% eye drops 1 drop(s), Eye-Both, 6 hourly
gentamicin 0.3% eye drops 1 drop(s), Eye-Both, 4 hourly
gentamicin 0.9% eye drops (prepared by pharmacy)
gentamicin 7.5 mg implant device
gentamicin 7.5 mg implant device 30 each, Intraosseous, ONCE, bone/soft tissue infection
gentamicin bladder irrigation
gentamicin bladder irrigation mg, Bladder Irrigation, ONCE, Other (specify in indication), Include dilution instructions in order comment
Gentamicin Level

#### CONCLUSIONS

The underlying mechanism of the error was 3.1 Selection errors when ordering

#### Example 5: Error not classified as a TRE

**Clinical error type:** Wrong dose

**Details:** *paracetamol 440 mg, QID, regular***.** Patient is has a weight in the 98.8th percentile for this age, with no height recorded. The weight 50th percentile for this age (5 years old) is 18 kg, giving a recommended dose of 270 mg. 440 mg gives a dose of 24.4 mg/kg.

| | |
|---|---|
| paracetamol<br>440 mg, Oral, QID, Analgesia, Review after 48 hours. | 440 mg @1251 |
| | 440 mg @1648 |

##### STEP 1: REVIEW ORDER IN THE CPOE

**Investigational question 1: Was the order based on a prebuilt order sentence?**

On investigation it was noted that the order was based on a prebuilt order sentence, as indicated by the inclusion of information such as the 'Drug Dosage Guidelines' embedded in the order sentence. These components were usually not completed by a prescriber when constructing an order without an order sentence.

| Details | |
|---|---|
| Target dose | 15 mg/kg |
| Actual dose | 15.17 mg/kg |
| Strength Dose | 440 |
| Strength Dose Unit | mg |
| Route of Administration | Oral |
| Frequency | QID |
| First Dose URGENCY | Routine |
| FIRST DOSE Date/Time | 20/05/2016 12:00 AEST |
| Indication | Analgesia |
| Special Instructions | Review after 48 hours. |
| Dose Limit | 90mg/kg/day (up to 4g) |
| Drug Dosage Guidelines | CHW: 15mg/kg/dose q4-6h |
| Next dose date and time | 20/05/2016 12:00 AEST |

**Investigational question 2: Was there information in the order as to how the dose had been determined?**

It was noted that the dose calculator was used to calculate the dose (as seen by the symbol next to the dose).

On accessing the dose calculator it was noted that no height had been recorded for the child, and the paracetamol dose was based on the child's actual weight, rather than ideal body weight as recommended by the hospital protocol for calculation of paracetamol doses.

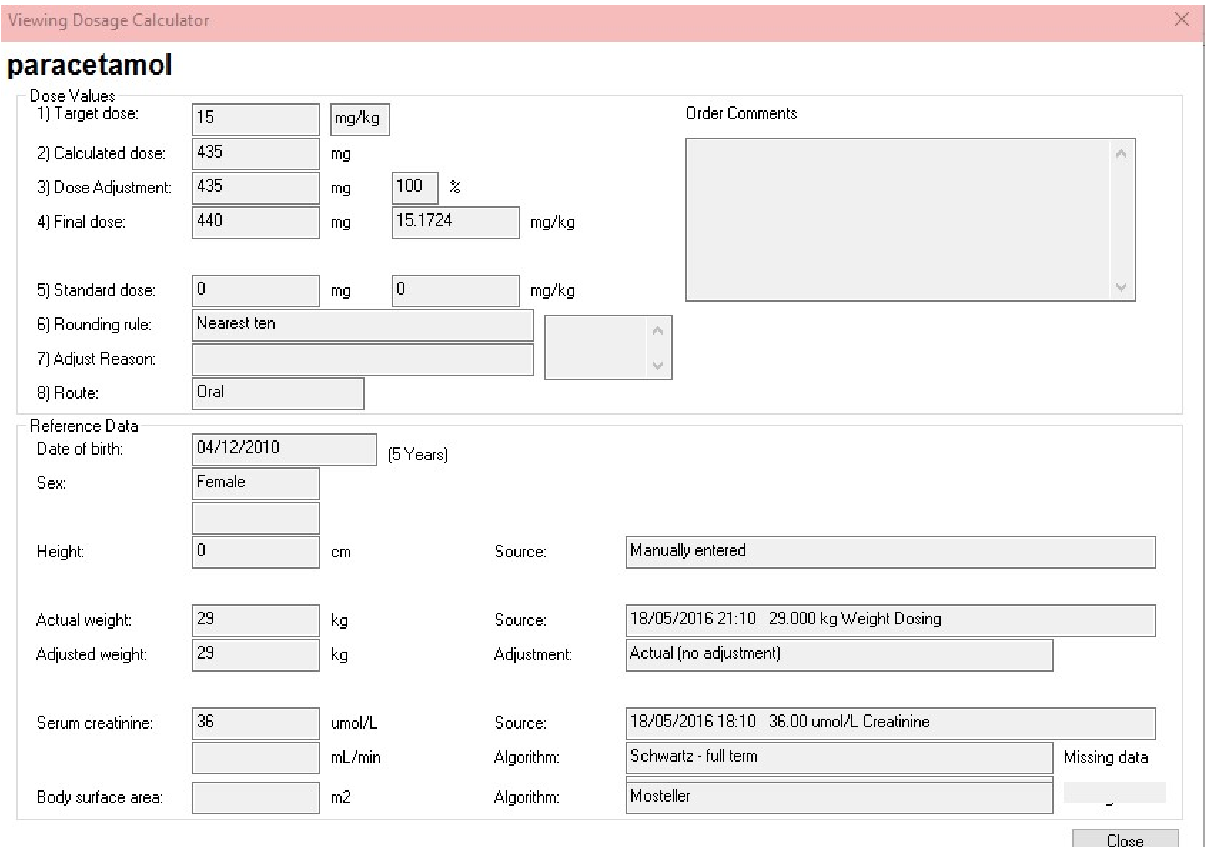
Viewing Dosage Calculator

**paracetamol**

Dose Values

1) Target dose: 15 mg/kg

2) Calculated dose: 435 mg

3) Dose Adjustment: 435 mg 100 %

4) Final dose: 440 mg 15.1724 mg/kg

5) Standard dose: 0 mg 0 mg/kg

6) Rounding rule: Nearest ten

7) Adjust Reason:

8) Route: Oral

Order Comments

Reference Data

Date of birth: 04/12/2010 (5 Years)

Sex: Female

Height: 0 cm Source: Manually entered

Actual weight: 29 kg Source: 18/05/2016 21:10 29.000 kg Weight Dosing

Adjusted weight: 29 kg Adjustment: Actual (no adjustment)

Serum creatinine: 36 umol/L Source: 18/05/2016 18:10 36.00 umol/L Creatinine

mL/min Algorithm: Schwartz - full term Missing data

Body surface area: m2 Algorithm: Mosteller

Close

**STEP 2: SIMULATE THE ERROR IN THE TEST ENVIRONMENT**

**Investigational question: Is there an error in the order sentence or template?**

In this scenario, there was no error in the build or programming of the order sentence. No further simulation of the error was required.

**CONCLUSIONS**

It was therefore determined that paracetamol overdose was not caused by the system, rather was an omission on the part of the prescriber in ensuring all the required patient parameters were used for correct calculation of the dose: the error is not technology-related.
