## Supplementary file 2 for "Understanding technology-related prescribing errors for system optimisation: the Technology-Related Error Mechanism (TREM) classification"

### SUPPLEMENTARY FILE 2: Technology-Related Error Mechanism (TREM) classification with examples

**Definition of a technology-related prescribing error:** An error where there is a high probability that the functionality or design of the technology contributed to the error, including errors arising from new work processes and changes in prescribing workflows that were implemented with the introduction of the technology. Excludes errors that the technology failed to prevent due to an absence of decision support unless existing decision support was not functioning as expected.

The underlying mechanism(s) of the technology related error (TRE) describe 'how' the error occurred (Table S1). More than one category of mechanism from the TREM classification may be associated with one TRE.

**Table S1: Major categories and sub-categories of the Technology-Related Error Mechanism (TREM) classification with examples**

| Major category | Subcategory | Examples |
|---|---|---|
| **1. Incorrect system configuration or system malfunction**<br><br>Errors within the system | 1.1 System malfunction<br><br>An isolated, usually temporary malfunction or fault. | • Dose decision support information for ibuprofen is temporarily displayed in the background of an individual order for paracetamol. |
| | 1.2 System contains incorrect order sentence* or other incorrect configuration | • Fentanyl order sentence was built incorrectly with the route 'intranasal-both' instead of 'intranasal', leading to an order with an unintentional double dose.<br>• Mupirocin nasal ointment and mupirocin ointment for topical use were incorrectly cross linked to the topical and nasal routes, respectively.<br>• System rounding rules within the dose calculator resulted in an order with an overdose or underdose. |
| | 1.3 Limitation in system functionality | • Search functionality not able to recognise medications when an alternate spelling was used, such as magnesium sulphate versus magnesium sulfate.<br>• When medication was ordered with the units of "mL" for a liquid requiring a mg/unit dose, no total dose was visible on the electronic medication administration record. |
| **2. Opening or using the wrong patient record** | | • Insulin was prescribed with the indication specified as type 1 diabetes mellitus in a patient with no history or recent diagnosis of diabetes, or any other indication for this medication. Medication was later cancelled. |

| | | |
|---|---|---|
| Errors that occur when prescriber uses an incorrect patient record.<br><br>May include mistyping a medical record number or name, selecting an incorrect patient from a list, inadvertently navigating to a previously opened record, or accessing a terminal that already has a record opened by another user. | | |
| **3. Selection errors**<br><br>Errors that occur when any element during prescribing is selected incorrectly from pre-programmed options presented by the system e.g. from a drop-down menu. | 3.1 Selection errors when ordering | • Prescriber selected phenoxymethylpenicillin instead of phenytoin from the medication drop-down menu.<br>• Prescriber selected an order sentence incorrectly from within an order set.⁑<br>• Prescriber selected heparinised saline instead of heparin.<br>• Prescriber selected an order sentence with the wrong form of methylprednisolone injection.<br>• Prescriber selected the wrong route (IV bolus) for vancomycin from drop-down menu.<br>• Prescriber selected the wrong dose form from a drop-down menu.<br>• Two similar items are selected from order set options creating therapeutic duplication e.g. morphine and fentanyl post-operative orders were both selected.⁑ |
| | 3.2 Selection errors when constructing or editing an order | • Prescriber selected an incorrect route, frequency or other element when editing an order sentence or constructing an order.<br>• Prescriber ordered pantoprazole 8mg/hour intra-articular daily, 200mg pantoprazole in 500mL sodium chloride 0.9%. The intra-articular route was selected instead of the intended intravenous infusion route.<br>• Prescriber ordered sodium chloride 0.9% via epidural route instead of intravenous route. |
| **4. Construction errors**<br><br>Errors that occur when constructing an order or typing free text, rather | | • A free text dose was prescribed with a typographical error in the units: ‘gm’ instead of ‘mg’, creating a 1000 x overdose.<br>• During construction of a free text order, a comment was inserted that did not match the rest of the order. |

| | | |
|---|---|---|
| than selecting from drop-down lists or editing order sentences. | | • Prescriber constructed an order for glyceryl trinitrate patch 10mg/24 hour, 1 patch in the morning, with order comment: 50mg/24 hour. The prescriber wanted the 50 mg patch which delivers a dose of 10mg/24 hours, not 5 patches.<br>• Prescriber entered patient's body mass index (15) into the dosing weight field, which was then used to calculate dose. Patient's actual weight was 6.36 kg.<br>• Prescriber typed dosing weight in as 13 g instead of 13 kg. System then used 13 g for the dose calculation.<br>• Prescriber inadvertently typed 6.48 kg instead of 4.68 kg in dosing weight field. |
| **5. Editing errors**<br>Errors that occur when editing (or not editing) a selected prepopulated order sentence or existing order (that are not selection errors or construction errors). | 5.1 Editing errors (general) | • Prescriber selected an ondansetron intravenous order sentence and changed the route to oral without removing "infuse over 15 minutes" from the order comments. |
| | 5.2 Editing errors that occur when using the dose calculator | • An error occurred when the prescriber rounded or manipulated the dose within the paediatric dose calculator resulting in an incorrect final dose. |
| | 5.3 Editing errors that occur when correcting a previous TRE | • An error in a previous order occurred relating to the dose calculator, leaving that order with an uncalculated mg/kg dose. An attempt was made to rectify the error by copying, not ceasing, the incorrect order leading to creation of a duplicate. |
| | 5.4 Editing errors that occur when the default time/date are not changed | • Order placed for insulin at 07:47 and given soon after. At 08:16 prescriber charted a regular ongoing pre-breakfast order intending for it to start the next day. The first dose however was automatically scheduled by the system for 09:00 that day, creating a duplicate with the order just given.<br>• Prescriber ordered fentanyl 50 microgram transdermal patch every three days at 13:46 on Monday. The task fell due immediately and at 13:46 every three days thereafter. However, the patient's current patch was due to be changed at 08:00 on Tuesday. |
| | 5.5 Editing errors that occur when misusing order actions on existing orders | • The prescriber intended to "cancel/discontinue" an order and instead inadvertently performed a "cancel/reorder" action causing a duplication. |
| **6. Errors that occur when using workflows that differ from a paper-based system** | 6.1 Updated medication profile, active workspace, or medication chart not viewed prior to ordering | • Errors that occurred when the prescriber failed to refresh the medication profile to allow viewing of current and most recent orders prior to creation of new orders.<br>• Prescriber inadvertently signed off on multiple unsigned draft orders (for example in a scratchpad†) for the same medication. |

| | | |
|---|---|---|
| | | • Medication already on the chart was prescribed using an order set.⁑ |
| | 6.2 Future order is not activated or a planned/pending future order or current activated order is not viewed | • Omalizumab was prescribed as a ‘future order’ for administration at a future admission, and as such was visible in the future orders section of the electronic chart, but not on the electronic medication administration record. On admission, a second prescriber charted omalizumab as an inpatient order, creating a duplicate with the existing future order. |
| | 6.3 Misuse of actions when ordering discharge or outpatient prescriptions, or when ordering from medication history or using medication reconciliation functionality | • Prescriber inadvertently ordered inpatient medication when attempting to order a discharge or outpatient prescription.<br>• Prescriber inadvertently or incorrectly ordered inpatient medication when attempting to reconcile medication history on admission, convert medication documented on admission to an active inpatient order, or during medication reconciliation process on discharge.<br>• Failure to edit incomplete or inaccurate medication history when converting to inpatient order. |
| | 6.4 Errors when using tasks and reminders | • An error that occurred in the use of a placeholder[‡] or ancillary task used to indicate the patient has medications prescribed on a paper pain chart.<br>• An order for warfarin was ceased, but the associated order for a warfarin check (INR prompt) remained active or vice versa. |
| | 6.5 Other<br><br>Other prescribing errors related to the new workflow required by the system | • Duplicate fluid orders that occur due to a change in fluid ordering workflow.<br>• Errors due to the change in the way to prescribe a medication with an unevenly split daily dose e.g. topiramate 25 mg morning and 50 mg night.<br>• Changes in workflow as a result of new pharmacy actions provided by the system e.g. a duplicate order created when pharmacy ‘rejected’ the order in the system, and prescriber allowed rejected order to remain active. |
| | 7.1 Errors occurring during initial system rollout (transition from paper to electronic) | • Incorrect cephalosporin was transcribed from paper into the electronic system e.g. cefotaxime vs ceftriaxone. |

| | | |
|---|---|---|
| **7. Contributing factor: use of hybrid systems**<br><br>Errors that occur when two different systems are used for prescribing including some prescribing remaining on paper medication charts or the use of different electronic systems.<br><br>Note: This category most commonly co-occurs with another mechanism. | 7.2 Errors occurring during downtime | • Errors related to the transition from the electronic system to paper charts and vice versa during system downtime. |
| | 7.3 Errors occurring when paper charts are used for some prescribing | • Opioid analgesics were prescribed in the electronic system creating duplication with patient-controlled analgesia prescribed on a paper pain chart. |
| | 7.4 Errors occurring when different electronic systems operate within the same hospital | • Incorrect transcription or reconciliation of orders on transition from intensive care using one electronic system to a general inpatient ward which used a different electronic system. |

Note: There may be more than one underlying mechanism for one prescribing error.
INR, international normalized ratio; TRE, technology-related error.
*An order sentence is a prewritten prescription sentence or prescription template which is based on the most common options prescribed for a medication, and often indication.
⁑An order set is a group of bundled standard items that are often ordered at the same time. In Cerner Powerchart™ this is termed a PowerPlan™.
†A scratchpad is an area of the system workflow that allows prescribers to create multiple draft prescriptions before sign-off.
‡A placeholder is an order that serves as a notification for an associated ancillary task or reminder.
